# Parkinson’s disease and multiple system atrophy are gateways to *RFC1*-related disorders

**DOI:** 10.1101/2025.08.18.25332961

**Authors:** V Delforge, G Coarelli, E Mutez, A-S Rolland, A Wissocq, A Labudeck, C Marzys, N Boucetta, L Buée, D Blum, A-R Marques, F Demurger, J-P Azulay, A Lanore, PREDISTIM study group, C Tesson, S Lesage, A Brice, D Grabli, D Devos, A Durr, V Huin

**Author notes:** Corresponding authors: DELFORGE Violette, Address: Permanent address: Inserm UMRS1172, ‘Alzheimer & Tauopathies’, Biserte building, Place de Verdun, 59045 Lille Cedex, France., HUIN Vincent, Address: Permanent address: Inserm UMRS1172, ‘Alzheimer & Tauopathies’, Biserte building, Place de Verdun, 59045 Lille Cedex, France. These authors contributed equally to this work (D Grabli, D Devos, A Durr).

## Abstract

Biallelic pathogenic expansions of the *RFC1* gene are the genetic cause of cerebellar ataxia, neuropathy, and bilateral vestibular areflexia syndrome. Sensory neuropathy is the most common symptom, but the clinical impairments and gateways to *RFC1*-related diseases are extremely variable. We genotyped patients with parkinsonism to test the hypothesis that this condition is another such gateway.

We screened four cohorts of patients with parkinsonism (*n* = 2037) for pathogenic expansions in the *RFC1* gene. In patients bearing two pathogenic expansions, we excluded the possibility of other pathogenic variants by exome sequencing. We detected 10/2037 (0.5%) biallelic (AAGGG)_n_ *RFC1* expansions. The initial diagnosis was Parkinson’s disease in five patients, multiple system atrophy in three and atypical parkinsonism in two.

Phenotypes perfectly mimicking Parkinson’s disease and multiple system atrophy defined according to the international diagnostic criteria may serve as gateways to some *RFC1*-related disorders. These results could modify the diagnostic and management of these two diseases. The diagnosis and follow-up of patients with parkinsonism should include searches for typical features evocative of a *RFC1*-relative disease, such as sensory neuropathy or chronic cough. Nerve conduction studies should be conducted in patients with unexplained symptoms of neuropathic pain or sensory neuropathy. We also suggest that *RFC1* screening should be performed in patients with atypical parkinsonism or multiple system atrophy with a long survival.

Trial Registration Information: The PREDISTIM cohort is registered with ClinicalTrials.gov: NCT02360683

**Graphical abstract:** 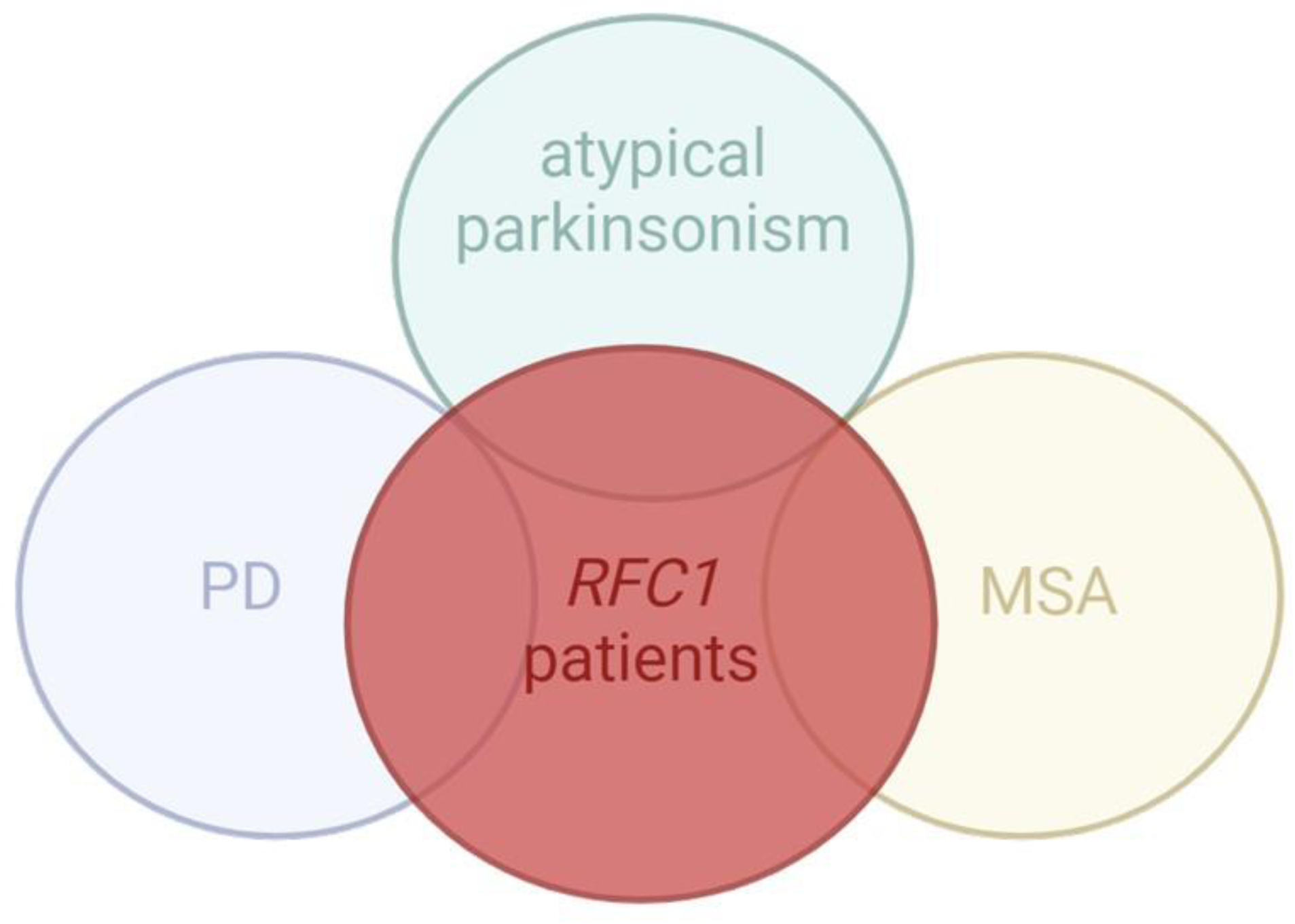

MSA = multiple system atrophy; PD = Parkinson’s disease; RFC1 = replication factor C subunit 1

## Introduction

In 2019, biallelic (AAGGG)_n_ repeat expansions in intron 2 of the *RFC1* gene were found to be the genetic cause of cerebellar ataxia, neuropathy, and bilateral vestibular areflexia syndrome (CANVAS) ^1,2^. The phenotypes associated with *RFC1* expansions are complex and involve multiple neurological symptoms. Chronic cough and sensory neuropathy are the most common gateways to *RFC1*-related diseases ^3^, but the inaugural symptoms are extremely variable. Rare patients may initially present with isolated hereditary sensory and autonomic neuropathy or isolated vestibulopathy.^4–8^ In most patients, these clinical impairments subsequently progress to more complex and multisystemic phenotypes, such as CANVAS ^9^. As the genetic cause of CANVAS was only recently discovered, it is unlikely that all the phenotypes and gateways to *RFC1*-related diseases have yet been identified.

Parkinsonism, which has three cardinal motor manifestations, is defined as bradykinesia combined with rest tremor, rigidity, or both ^10^. It is observed in neurodegenerative diseases, such as Parkinson’s disease (PD), the most frequent cause of parkinsonism, and in atypical Parkinsonian disorders, which can have multiple etiologies, including multiple system atrophy (MSA), progressive supranuclear palsy (PSP), corticobasal syndrome and dementia with Lewy bodies. We found that 4/38 (10.5%) patients in a CANVAS cohort had parkinsonism, a frequency 10 times higher than would be expected in the general population matched for age.^11^ Moreover, parkinsonism has been described in other studies of patients with *RFC1-*related diseases and is now considered part of the extended phenotype of *RFC1* disorder.^6,9,11–18^ Two studies by a Finnish team reported 3/559 and 3/273 patients with biallelic pathogenic expansions in cohorts of PD and early-onset PD patients, respectively.^19,20^ Some of these patients had clinical signs not usually observed in classical PD, such as vestibular dysfunction, sensory neuropathy, dysautonomia, and/or chronic cough. Jerez *et al.*^21^ recently described four patients with biallelic pathogenic expansions in a cohort of 1609 patients with PD. Liu *et al.*^22^ reported two other *RFC1* patients in a cohort of 1445 patients with parkinsonism.

The similarity of the clinical symptoms of CANVAS and MSA of the cerebellar type (MSA-c) have led to searches for *RFC1* expansions in MSA cohorts. The clinical symptoms common to these two conditions include dysautonomia, cerebellar ataxia, parkinsonism and cognitive impairment. Some studies found no pathogenic repeat expansions in MSA cohorts,^23–25^ whereas others reported 3/207, 3/44 or 3/282 MSA-like patients with biallelic (AAGGG)_n_ expansions.^26–28^ However, some of these patients had additional symptoms typical of CANVAs, such as vestibular impairment or chronic cough. Moreover, at least five of the nine MSA-like patients previously reported to have biallelic *RFC1* expansions had sensory neuronopathy, as demonstrated by nerve conduction studies (NCS).^26,28^ The overlap of symptoms makes it difficult to differentiate CANVAS from MSA-c and only certain “red flags” for MSA-c, such as sensory neuropathy are useful for differential diagnosis. It has not yet been demonstrated that *RFC1* biallelic expansion can be misdiagnosed as definite or probable MSA-c according to the international diagnosis criteria.^29^ There is, therefore, no consensus on whether *RFC1* screening should be performed in patients suffering from MSA-c.

We hypothesised that parkinsonism may be a gateway to *RFC1*-related disorders. In that case, it would be important to determine what differentiate *RFC1* patients with parkinsonism as their first symptom from the other parkinsonism patients. Indeed, these *RFC1* patients with parkinsonism may display specific clinical impairments and/or different evolution that may need other management of their disease compared to the classical PD or MSA patients. To answer these questions, we screened four cohorts of patients with parkinsonism for pathogenic expansions in the *RFC1* gene, and we report here the detailed phenotype of biallelic expansion carriers.

## Materials and methods

### Patients

Four cohorts of patients with parkinsonism were analysed in this study. The PARK cohort consisted of 702 affected patients from 686 families with parkinsonism or with Parkinson’s disease of strongly suspected genetic origin due to the early onset of the disease (<50 years) and/or a positive family history. Between June 2007 and December 2022, neurologists or geneticists collected peripheral blood in tubes containing ethylenediaminetetraacetic acid, which were then sent to the Laboratory of Neurobiology of Lille University Hospital for molecular diagnosis. The patients were mostly seen at French university and regional hospitals. We had previously screened all samples for the most frequent pathogenic variants causing parkinsonism by multiplex ligation-dependent probe amplification (Probe mixes P051 and P052; MRC Holland, Amsterdam, The Netherlands).^30^ We excluded DNA samples from minors and patients with an established molecular diagnosis.

The second cohort of 773 unrelated patients came from an ancillary study of the Predictive Factors and Subthalamic Stimulation in Parkinson’s disease study (PREDI-STIM) https://clinicaltrials.gov/ct2/show/NCT02360683?term=predi+stim&draw=2&rank=1. This study is a broad French prospective multicentre study with the standardised collection of clinical, imaging and genetics data for PD patients with advanced disease and candidates for deep brain stimulation (DBS).

The SPD cohort contained 368 patients from 360 different families. Those patients have been selected based on the association of Parkinson’s disease with dysautonomia. The MSA-c cohort comported 194 patients with a clinical diagnostic of “probable” multiple system atrophy of cerebellar type from 194 different families. The patients from the SPD and MSA-c cohorts were included in the SPATAX study (NCT00140829, CPP Ile de France VI, AOM10094) and BIOMOV study (NCT05034172, CPP SUD-EST IV, 2021-A00989-32), followed at both the Pitié-Salpêtrière Hospital in Paris and/or Paris Brain Institute.

Clinical diagnoses of PD and MSA in the patients and affected relatives were reviewed according to the diagnostic criteria for PD ^10^ and multiple system atrophy ^29^.

### Molecular analysis

Screening for the pathogenic expansion (AAGGG)_n_ was performed by polymerase chain reaction (PCR). We first performed duplex PCR with fluorescent primers as previously described ^31^ and repeat-primed-PCR (RP-PCR) targeting the pathogenic motif (AAGGG)_n_ as described by Cortese *et al*.^1^ The duplex PCR was used to detect the presence of two expanded alleles (absence of amplification), whereas the RP-PCR (AAGGG)_n_ was used to detect the presence of at least one pathogenic expansion. In patients with no amplification on duplex PCR and/or with an expansion (AAGGG)_n_ detected by the first RP-PCR, we performed two other RP-PCRs targeting the frequent (AAAAG)_n_ and (AAAGG)n expansions.^1^ We investigated the possible presence of the pathogenic motif in both alleles by performing Southern blots on long-range PCR products, as previously described.^11^

In patients bearing two pathogenic expansions (AAGGG)_n_ at the *RFC1* locus, we excluded the presence of any other pathogenic variant that might explain the phenotype by exome sequencing on a NovaSeq6000™ instrument. We used DRAGEN™ software (Illumina, San Diego, California, USA). Exon capture and library preparation were performed with the TrueSeq DNA Exome S4 kit (Illumina, San Diego, CA, USA), according to the manufacturer’s instructions. Pooled libraries were sequenced by paired-end, 150-cycle chemistry on an Illumina NovaSeq6000 (Illumina, San Diego, CA, USA). The sequence data were aligned with the hg38 assembly version of the human genome, with BWA v0.7.17. Variant calling, joint genotyping, and recalibration were performed with GATK v3.7. Variant annotation was performed with Variant Effect Predictor (https://www.ensembl.org/info/docs/tools/vep/index.html) and Dragen software (Illumina, San Diego, CA, USA). SIFT, Polyphen2 HumDiv, Polyphen2 HumVar, FATHMM, AlphaMissense, REVEL, ClinPred, Meta SVM, Meta LR, and Mistic predictions of deleteriousness were obtained with Variant Effect Predictor (https://www.ensembl.org/info/docs/tools/vep/index.html). We used DRAGEN CNV Baseline Builder (Illumina, San Diego, CA, USA) to analyse structural variations.

### Ethics

Patients or their legal caregivers gave their written informed consent for genetic testing and publication of the relevant findings. All clinical examinations were performed in accordance with the Declaration of Helsinki (2013). The studies were approved by the competent ethics committees in accordance with French ethics regulation: The study is registered with the “Commission Nationale de l’Informatique et des Libertés” (CNIL) numbers 726/2020 and 1418/2022; “Nord Ouest-IV” Ethics Committee (IDRCB N°: 2013 A0019342), “Paris Necker” Ethics Committee (RBM 03-48 and RBM 01-29), and ClinicalTrials.gov (NCT02360683). The SPATAX study (NCT00140829, CPP Ile de France VI, AOM10094) and the BIOMOV study (NCT05034172, CPP SUD-EST IV, 2021-A00989-32) were also registered separately.

## Results

### Demographics

All 2037 patients with parkinsonism were screened for *RFC1* expansions. Details of the four cohorts are provided in Supplementary data.

### Genetic screening

In total, 164/2037 individuals (8%) were heterozygous for the pathogenic expansion (AAGGG)_n_ and 1863/2037 (91.5%) did not have the pathogenic expansion in either allele. The details for each cohort are provided in Supplementary Figure S1. In 10/2037 individuals (0.5%), we detected biallelic (AAGGG)_n_ *RFC1* expansions. The details for each patient with biallelic (AAGGG)_n_ expansions are provided in Table 1.

**Table 1.** Characteristics of the 10 *RFC1* patients with parkinsonism. ^1^ Stridor, frequent sighs; ^2^ Hyperkinetic movement, but not including levodopa-induced dyskinesia; Y: Yes; N: No; Na: not available; NC: not concerned; RBD: REM sleep behavior disorder; M: Man; W: Woman; MoCA: Montreal Cognitive Assessment; MMSE: Mini Mental State Examination; DBS: deep brain stimulation; PD: Parkinson’s disease; MSA-c: multiple system atrophy of the cerebellar type; CANVAS: Cerebellar ataxia, neuropathy, and bilateral vestibular areflexia syndrome; UPDRS: Unified Parkinson Disease Rating Scale; NCS: Nerve Conduction Study; DAT-scan: [123I]-ioflupane dopamine-transporter single-photon emission computerised tomography; MRI: magnetic resolution imaging; PREDISTIM: Predictive Factors and Subthalamic Stimulation in Parkinson’s disease study.

| Cohort | Code Patient | Initial diagnosis (parkinsonism) | Demographic |  |  |  |
| --- | --- | --- | --- | --- | --- | --- |
|  |  |  | Sex | Age at onset<br>in years | Age at first<br>walking aid | Age at last<br>examination |
| MSAc | AAD-SAL-ROB-769-007 | probable MSA-c | M | 41-45 | 51-55 | 56-60 |
| MSAc | SAL-LAU-399-139 | probable MSA-c | W | 51-55 | *na* | 56-60 |
| MSAc | SAL-MAR-399-358 | probable MSA-c | M | 41-45 | *na* | 61-65 |
| PREDISTIM | 050006109 | PD | W | 56-60 | 61-65 | 71-75 |
| PREDISTIM | 060053109 | PD | M | 51-55 | *na* | 66-70 |
| Atypical PD | SPD-AIX-RIC-550-004 | PD | W | 46-50 | *na* | 61-65 |
| Inherited PD | PARK182 | PD | M | 46-50 | NC | 61-65 |
| Inherited PD | PARK607 | PD | M | 56-60 | 66-70 | 71-75 |
| Inherited PD | PARK194 | atypical PD | M | 61-65 | 71-75 | 71-75 |
| Inherited PD | PARK728 | atypical PD | M | 56-60 | *na* | *na* |

|  |  | Parkinsonism features |  |  |  |  |  |  |  |
| --- | --- | --- | --- | --- | --- | --- | --- | --- | --- |
| Age at death<br>in years | Cause of<br>death | Akinesia<br>Bradykinesia | Rigidity | Resting<br>tremor | Asymmetric<br>Symetric | Levodopa-induced | Dopa-sensitive | UPDRS III<br>On/Off | Marked on/off<br>fluctuations end- |
| 61-65 | na | Y | Y | Y | Symetric | na | N | na | na |
| 61-65 | na | Y | N | Y | Symetric | na | na | na | na |
| 66-70 | na | Y | Y | N | Symetric | na | na | na | na |
| 71-75 | Fall | Y | Y | Y | Asymmetric | Y | Y | 11.5/34.5 | Y |
| na | na | Y | Y | N | Asymmetric | N | Y | 9 | Y |
| 66-70 | na | Y | Y | Y | Asymmetric | Y | Y | 24/49 | Y |
| Alive | na | Y | Y | Y | Asymmetric | Y | Y | 15/23 | Y |
| 71-75 | Aspiration | Y | Y | Y | Asymmetric | Y | Y | 14/48 at 61-65, 29/46 | Y |
| 71-75 | na | Y | Y | N | Symetric | N | N | 21/25 | N |
| 71-75 | na | Y | Y | Y | Symetric | na | Y | na | na |

Whole-exome sequencing on the 10 patients with biallelic pathogenic *RFC1* expansions identified no pathogenic variant that could have caused inherited parkinsonism. However, patient SPD-AIX-550-4 was heterozygous for a pathogenic variant in exon 7 of the *GBA1* gene (NM_000157.4:c.876del, p.(Glu293Asnfs*11)). This variant was not sufficient to account for disease in the patient, but may constitute a genetic risk factor for PD.^32^

### PARK cohort

In this cohort, 4/702 (0.57%) patients had biallelic pathogenic expansions. Parkinsonism occurred at a median age at onset of 58 years (interquartile range (IQR): 54.5 - 61.5) and all four patients were men. The inaugural symptom was parkinsonism in 3/4 (75%) patients. Two patients had an initial diagnosis of PD because they were dopamine-sensitive and had no exclusion criteria for PD. Both had dysautonomia and REM sleep behaviour disorder and were treated with clozapine for hallucinations and with a duodenal infusion of levodopa/carbidopa gel (Duodopa®). Both these patients later developed a sensory neuropathy, as determined by NCS and, the presence of neuropathic pain. One patient had a chronic cough three years after parkinsonism onset. The other two patients had atypical parkinsonism that was difficult to classify according to current criteria. One had a balance disorder as the first symptom between the ages of 61 and 65 years. He developed dysarthria during this same period and symmetric levodopa-resistant parkinsonism associated with a pyramidal syndrome and severe cognitive impairment between his 66 and 70 years. Brain MRI results were consistent with MSA-p including a putamen hyposignal, but rapid cognitive deterioration was observed, with a frontal syndrome and dementia in these years, followed by progression to supranuclear ophthalmoplegia and retrocollis between the ages of 71 and 75 years, with symptoms potentially also compatible with PSP. A sensory neuropathy was fortuitously discovered during NCS for the diagnosis of carpal tunnel syndrome. A review of the patient’s medical history revealed a chronic cough that appeared between the ages of 51 and 55 years. The final patient had symmetric, levodopa-sensitive parkinsonism beginning between 56 and 60 years. PSP was also suspected, due to the rapidity of cognitive impairment and vertical ophthalmoplegia. This patient had pyramidal syndrome, mild cerebellar ataxia and cognitive impairment.

### PREDISTIM cohort

In this cohort of patients with evolved PD, 2/773 (0.26%) had biallelic pathogenic *RFC1* repeat expansions. These two patients were diagnosed with PD in their 50’s. One was male and the other was female. Both were dopamine-sensitive with marked on/off fluctuations. Their PD followed a normal course, with no CANVAS-like symptoms. One displayed a rapid progression of motor symptoms, dysphagia and hyperkinetic movements.

### SPD cohort

One patient with biallelic pathogenic *RFC1* repeat expansions was identified among the 368 patients of this cohort (0.27%). She was diagnosed with PD at the end of her 50’s, was dopa-sensitive, with asymmetric disease and marked on/off fluctuations. She had dysautonomia, mainly orthostatic hypotension, and dysarthria. She was treated with DBS since between the ages of 56 and 60 years and died at the end of her 60’s.

### MSA-c cohort

The frequency of homozygous carriers was highest in the MSA-c cohort with 3/194 (1.55%) patients positive for biallelic *RFC1* pathogenic expansions. The median age at onset for *RFC1* patients was 45 years (IQR: 45 – 48 years). All three patients were Caucasian and two were male. Their parkinsonism symptoms were symmetric and consisted of bradykinesia and rigidity in two cases and bradykinesia and resting tremor in the third. All three patients had dysautonomia, dysphagia, pyramidal signs, and cerebellar ataxia with cerebellar and brainstem atrophy on brain MRI. Interestingly, none of these patients presented clinical signs of sensory neuropathy. However, two patients had a sensory neuropathy detected by NCS. One patient had rapidly progressing motor symptoms, a sleep disorder and impairment in cognitive tests. These three patients with *RFC1* patients had a diagnosis of probable MSA of the cerebellar type according to international criteria.^29^ However, all three patients had features unusual for classical MSA. Indeed, all patients had a slow progression of the disease evidenced by an improved survival. The median of disease duration until death was 16 years and one patient died 25 years after the disease onset. After excluding these 3 *RFC1* subjects, the patients from the MSA-c cohort had a median survival of 9 years [7 to 11] (n = 127). Relative to the other patients from this cohort, the three *RFC1*-MSA-c patients had a longer survival (p = 0.047; Wilcoxon rank sum test with continuity correction).

## Discussion

In this study, we screened patients with parkinsonism and found that 10/2037 (0.5%) had biallelic pathogenic *RFC1* expansions. According to the allelic frequency in the general population (0.045),^33^ we would expect no more than 4.1 individuals per 2037 people in the general population carrying biallelic *RFC1* expansions; here, the frequency of such expansions was more than double the expected value. Given (i) the late onset in *RFC1*-related disorders, (ii) the high prevalence of parkinsonism (and especially PD) in the general population and (iii) the frequency of heterozygosity for pathogenic (AAGGG)_n_ expansions in Caucasians, we cannot exclude the possibility that some of the 10 patients with biallelic pathogenic *RFC1* expansions reported here had two different and unrelated neurodegenerative diseases. This is the case for the three PD patients without unusual disease progression treated with DBS, in particular. In these specific patients, it is difficult to assess the causality of *RFC1* pathogenic expansion in the development of their disease. However, these patients might later express some symptoms from the CANVAS-spectrum. The other *RFC1* patients reported here have a more complex phenotype and/or later progressed from typical PD or MSA to a neurological disease with symptoms resembling those of *RFC1*-related disorders. These results suggest that the parkinsonism in our patients may not be a comorbid condition in some cases.

We revealed biallelic pathogenic *RFC1* expansions in five patients diagnosed with PD who had been treated for several years. It was not possible to distinguish their disease from PD during the first years after onset in any of these patients. Three patients were treated by DBS and the other two had duodenal infusion of levodopa/carbidopa gel. Two of our five PD patients with biallelic pathogenic *RFC1* expansions developed chronic cough and/or sensory neuropathy and neuropathic pain after the diagnosis of PD. In previous reports, patients with *RFC1* expansions diagnosed with PD also had signs of polyneuropathy, chronic cough or vestibular impairment.^19,20^ CANVAS spectrum-related clinical impairment was not assessed on another study,^21^ and was absent in two PD patients from the study by Liu *et al.*^22^ Only 3/17 patients with *RFC1*-PD-like disease reported here and in previous studies underwent NCS (Supplementary Table S1). These observations suggest that a PD diagnosis according to the international diagnosis criteria can serve as a gateway to some *RFC1*-related disorders. Usual hallmarks of *RFC1*-related diseases can be overlooked in the context of PD. Indeed, NCS are not part of the usual investigation in PD, a mild sensory neuropathy can be easily missed, and chronic cough is not a symptom reported as a red flag for PD diagnosis.

Our findings suggest that changes are required in the management of patients with suspected PD. The diagnosis and follow-up of these patients should include searches for typical features evocative of CANVAS, such as sensory neuropathy or chronic cough. NCS should be conducted in patients with symptoms of neuropathic pain or sensory neuropathy, after the elimination of vitamin deficiency linked to levodopa treatment. It may be of interest to search for vestibular abnormalities too, although no such abnormalities were observed here. Our findings suggest that it is also of interest to ask for chronic cough during medical interview. Finally, further studies are warranted to define the role of systematic NCS in PD patients as a means of detecting subclinical sensory neuropathy, which may constitute an additional cause of handicap in these patients, leading to falls during disease progression that require appropriate care.

Three patients with *RFC1* expansions in our cohorts were diagnosed with MSA-c and fulfilled the criteria for probable diagnosis of this disease. Strikingly, these patients had a clinical phenotype very similar to that of classical MSA-c, including supportive features, such as stridor, early bulbar dysfunction and pyramidal syndrome. None of these patients had a history of chronic cough, but it should be borne in mind that this is a retrospective study and that questions about chronic cough are not part of the usual medical interview in patients with MSA. None of these patients complained of sensory neuropathy symptoms, but two were found to have sensory neuropathy on NCS. Two patients suffered from neuropathic pain. Again, these results highlight the potentially value of performing NCS in patients with MSA.

The involvement of *RFC1* expansions in MSA remains a controversial issue, with some studies finding no link.^23–25^ Three previous studies reported a total of nine patients with biallelic pathogenic *RFC1* expansions who had been diagnosed with possible or probable MSA.^26–28^ Only the phenotypes of six MSA patients were reported and in five of these six patients it was possible to distinguish between the patient’s condition and typical MSA as these patients also suffered from sensory neuropathy and/or vestibular impairment and/or chronic cough. The phenotype of the sixth patient was not investigated in detail (Supplementary Table S2). It is clear from these and our own studies that *RFC1* expansions can cause a phenotype mimicking MSA. We suggest that screening for *RFC1* expansions should be performed in patients with MSA, particularly in cases of MSA-c with long survival. Interestingly, survival seems to be greater for the *RFC1*-MSA-like patients in our study than for those with typical MSA. Indeed, the median survival of our three patients was 16 years whereas the median survival for MSA patients in published studies is less than 10 years.^34^ This finding contrasts with the results for “patient V” studied by Wan *et al.*^27^ who died four years after disease onset. However, the molecular diagnosis for this patient did not include Southern blotting as the patient died before this test could be performed. Prospective follow-up of these previously reported *RFC1*-MSA-like patients would be useful, to determine whether they do indeed have a better survival. If this should prove to be the case, then the genetic diagnostic of *RFC1* mutations in patients with MSA would be of the utmost importance to differentiate these patients from those with classical MSA patients and to provide a more accurate prognosis.

We report two other patients with atypical parkinsonism presenting some features of PSP, including supranuclear palsy, related retrocollis and dementia with frontal syndrome. Traschütz *et al.*^3^ suggested a possible overlap with PSP in their report of six patients with *RFC1*-related disorders co-occurring with bradykinesia with postural instability and a slowing of vertical saccades. Similar studies of *RFC1* screening in cohorts of PSP patients are warranted to confirm or infirm this new association.

This study has several limitations. Firstly, it was retrospective and it is therefore possible that the available information is incomplete. Some symptoms, such as chronic cough, may have been present in some patients but overlooked by neurologists. Secondly, our cohorts include mostly Caucasians, which may represent a bias. Further studies involving patients of different ethnicities and from different countries of origin are required to determine whether our results are applicable to all patients with *RFC1* expansions and parkinsonism, or whether they need to be interpreted in a specific genetic and/or environmental context. Thirdly, given the overrepresentation of Caucasians among our patients, we did not screen for other pathogenic motifs, such as (ACAGG)_n_, more frequently observed in Asian populations. Lastly, we do not have any neuropathological examination of the *RFC1* patients reported here, especially the patients with MSA-like phenotype. In previous studies, we and other teams reported the presence of synuclein aggregation and Lewy bodies in the brain of *RFC1* patients.^11,35^ It would have been interesting to search in these MSA-like patients if there was any synuclein aggregation too. In such case, *RFC1* pathogenic expansions would be one of the rare variant associated with different types of synucleopathies. Screening of *RFC1* expansions in cohorts of patients with other synucleinopathies such as MSA-p or dementia with lewy bodies would be of interest. This raises too the question of the putative link between *RFC1* and alpha-synuclein.

In 2025, Liu *et al.*^22^ reported the long-read sequencing of novel repeat configurations at the *RFC1* locus in two patients with *RFC1*-related PD. Once enough patients have been identified, it might be interesting to sequence the *RFC1* expansions in our patients with parkinsonism and compare them to those in patients with other phenotypes to look for unusual conformations or lengths that might explain the different phenotypes in *RFC1*-related disorders. Other diseases due to expansions in the *NOTCH2NLC*^36–38^ and *FGF14*^39–41^ genes have recently been discovered and associated with PD or MSA. We expect that this work will be followed by other reports defining new genetic causes of PD-like and/or MSA-like phenotypes.

In conclusion, we report the diagnosis of PD and MSA as a gateway to *RFC1*-related disease and expand the phenotype of these genetic disorders. Knowledge and recognition of an *RFC1*-related disorder in patients presenting with a PD or MSA phenotype will be essential for the diagnosis, prognosis and specific care of these rare patients. The search for symptoms of sensory neuropathy, neuropathic pain or chronic cough should be systematic in all patients with parkinsonism at the time of diagnosis and during follow-up. A genetic testing of *RFC1* expansions may also be considered in MSA patients with increased survival. Further studies are warranted to define the role of NCS in these diseases, which may increase with subsequent descriptions of new genetic causes of parkinsonism.

## Data Availability

The data are available from the corresponding author upon reasonable request.

## Acknowledgements

We thank all the patients and their relatives for participating in this study. We thank the French clinical research network NS-Park/F-CRIN and the Fédération de la Recherche Clinique du CHU de Lille for support (with Anne-Sophie Rolland, Alain Duhamel, Maeva Kheng, Julien Labreuche, Dominique Deplanque, Nolwen Dautrevaux, Victor Laugeais, Morgane Coeffet, Maxime Caillier, Aymen Aouni, Pauline Guyon, Francine Niset, Valérie Santraine, Marie Pleuvret, Julie Moutarde and Laetitia Thibault). We thank Dr. Sylvie Forlani and Ludmila Jornéa (Sorbonne Université, Paris Brain Institute, DNA/Cell Bank).

## Supplementary data

### Details of the four cohorts studied

#### 1) PARK cohort

This cohort includes 702 patients: 400 men (57%) and 302 women (43%). Median age at onset was 47 years [IQR: 40 to 57 years]. Most patients were Caucasian (660/702, 94%). The samples were sent to the Laboratory of Neurobiology of Lille University Hospital for molecular diagnosis, because of (i) early onset (<50 years) in 251 (35.8%), (ii) family history of parkinsonism in 149 (21.2%), or (iii) both in 302 (43%). The In total, 662 of the patients (94.3%) were diagnosed with PD.

#### 2) PREDISTIM cohort

This study is an ancillary analysis of data collected from the PREDISTIM cohort, an ongoing prospective multicentre cohort (Protocol 2013-A00193-42) sponsored by Lille University Hospital, conducted at 17 expert centres for PD from the French clinical research network (NS-Park/F-CRIN). The principal objective of this cohort study was the identification of factors predictive of the therapeutic response to STN-DBS in terms of long-term quality of life. Briefly, patients undergoing STN-DBS at each of the participating centres were consecutively included into the study between November 2013 and September 2019. The inclusion criteria were a diagnosis of PD according to the UK Parkinson’s Disease Brain Bank, a disease duration ≥5 years, age between 18 and 75 years, and an indication for STN-DBS. Exclusion criteria included atypical parkinsonism, severe cognitive impairment, severe psychiatric disorders, acute levodopa motor response <30%, and contraindications for surgery. Clinical data are collected at baseline and then one, three and five years post-surgery. The cohort includes 773 PD patients: 491 men (63.5%) and 282 women (36.5%). At the time of the study, the median age of the patients was 63 years [interquartile range (IQR): 58 to 67 years]. Median age at onset was 53 years [IQR: 50 to 55 years]. Most patients were Caucasian (712/773, 92.1%).

#### 3) MSA-c

The cohort includes 194 patients: 116 men (59.8%) and 78 women (40.2%). The median age at onset was 54 years [IQR: 49 to 60]. Most patients were Caucasian (187/197, 94.9%). The median age at last examination was 60 years [IQR: 54 to 66]. The median age at death was 64 years [IQR: 61.25 to 69.5], for a median disease duration of 10 [IQR: 6 to 11].

**Supplementary Figure S1.**
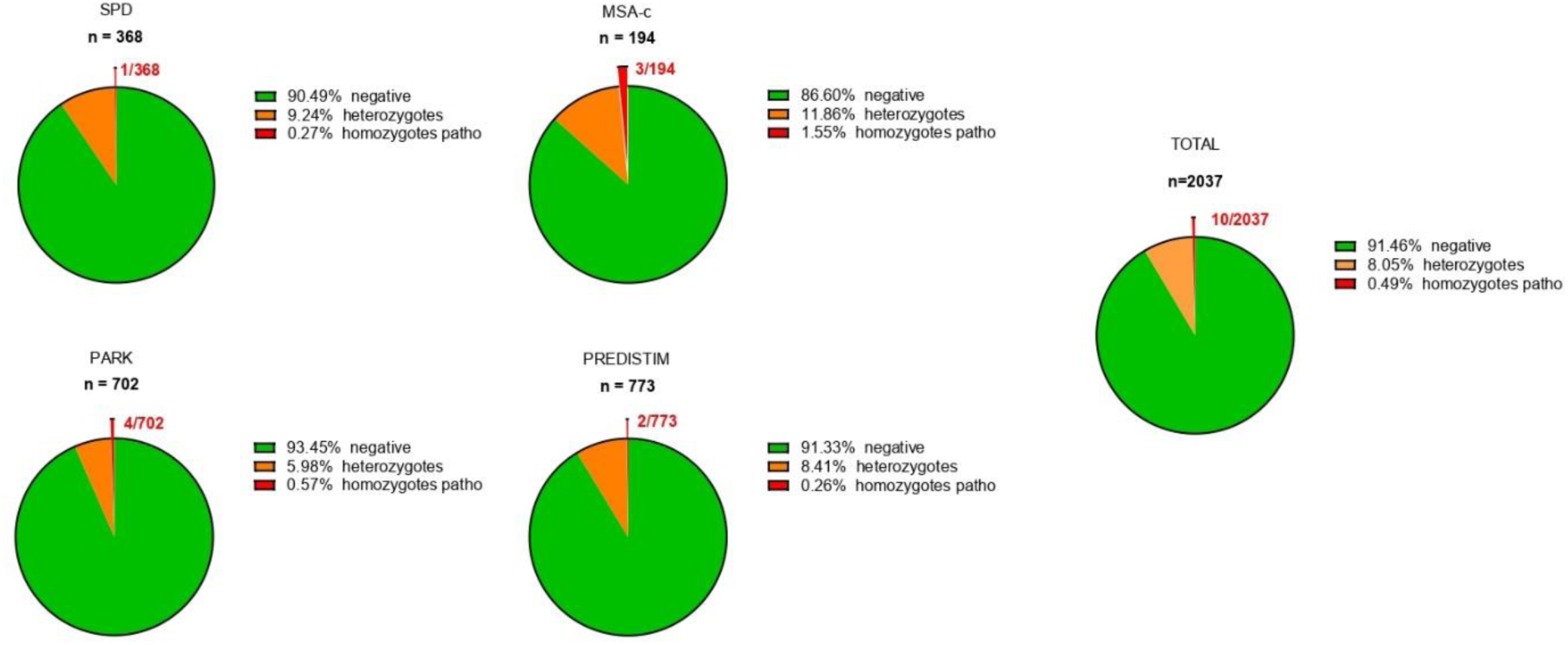
Results of *RFC1* screening in the four cohorts. Results of the screening of patients from each cohort, expressed as percentages. The percentage of heterozygotes was highest in the MSA-c cohort. MSA-c: multiple system atrophy of the cerebellar type; PREDISTIM: Predictive Factors and Subthalamic Stimulation in Parkinson’s disease study; Negative: no *RFC1* expansions; Heterozygotes: *RFC1* expansions (AAGGG)_n_ in one allele only; Homozygotes patho: *RFC1* expansions (AAGGG)_n_ in both alleles.

**Supplementary Table S1.** Comparison of the characteristics of our five *RFC1* patients diagnosed with PD with those of published cases. Y: Yes; N: No; na: not available; NC: not concerned; M: Man; W: Woman; MoCA: Montreal Cognitive Assessment; MMSE: Mini Mental State Examination; DBS: deep brain stimulation; PD: Parkinson’s disease; MSA-c: multiple system atrophy of the cerebellar type; CANVAS: cerebellar ataxia, neuropathy, and bilateral vestibular areflexia syndrome; UPDRS: Unified Parkinson Disease Rating Scale; DAT-scan: [123I]-ioflupane dopamine-transporter single-photon emission computerised tomography; MRI: magnetic resolution imaging; PREDISTIM: Predictive Factors and Subthalamic Stimulation in Parkinson’s disease study; MIBG: meta-iodobenzylguanidine.

| Inclusion/Exclusion criteria for PD |  |  |  |  |  |  |  |
| --- | --- | --- | --- | --- | --- | --- | --- |
| Rapid progression of motor | Rapid progression of cognitive | Cognition impairment | Neurocognitive testing | Early bulbar dysfunction | Inspiratory respiratory | Severe autonomic failure | Anterocollis or limb |
| Y | N | Y | *na* | Y | Y | Y | N |
| N | N | *na* | *na* | Y | *na* | Y | *na* |
| N | N | *na* | *na* | Y | *na* | Y | *na* |
| N | N | N | MoCA: 28/30 at 66-70 | N | N | N | N |
| N | N | N | MoCA: 28/30 at age 66-70 | N | N | N | N |
| N | N | N | *na* | N | N | N | *na* |
| N | N | Y | Mattis: 135/144 at 46-50, MMSE: 26/30 at | N | N | N | N |
| N | N | Y | MoCA: 23/30 at 61-65 | N | N | N | N |
| N | Y | Y | MMSE 23/30 at 66-70, Mattis 98/140 at 66- | Y | N | N | N |
| *na* | *na* | Y | *na* | *na* | *na* | *na* | *na* |

axia, neuropathy, and bilateral vestibular areflexia syndrome; UPDRS: Unified Parkinson Disease Rating Scale; NCS: Nerve Conduction Study; DAT-scan: [<sup>123</sup>I]-ioflupane dopamine-transporter single-photon emiss

**Supplementary Table S2.** Comparison of the characteristics of our three MSA patients with *RFC1* expansions with those of published cases. ^1^ Stridor, frequent sighs; ^2^ Hyperkinetic movement, but not including levodopa-induced dyskinesia; Y: Yes; N: No; na: not available; M: Man; W: Woman; MoCA: Montreal Cognitive Assessment; MMSE: Mini Mental State Examination; DBS: deep brain stimulation; PD: Parkinson’s disease; MSA-c: multiple system atrophy of the cerebellar type; MSA-p: multiple system atrophy of the parkinsonian type; CANVAS: cerebellar ataxia, neuropathy, and bilateral vestibular areflexia syndrome; RBD: REM sleep behavior disorder; DAT-scan: [123I]-ioflupane dopamine-transporter single-photon emission computerised tomography; MRI: magnetic resolution imaging; PREDI-STIM: Predictive Factors and Subthalamic Stimulation in Parkinson’s disease study; MIBG: meta-iodobenzylguanidine.

|  | Clinical features in common between PD or MSA or CANVAS-spectrum |  |  |  |  |  | Other CAN |  |
| --- | --- | --- | --- | --- | --- | --- | --- | --- |
| Pyramidal syndrome | Dysautonomia | Oculomotor abnormalities | Lower motor neuron signs | Sleep disorders | Cerebellar ataxia | Hyperkinetic movements<sup>2</sup> | Sensory neuropathy | Vestibular areflexia |
| Y | Y | Saccadic pursuit | N | Y | Y | N | N | na |
| Y | Y | Saccadic pursuit | N | na | Y | N | N | na |
| Y | Y | Saccadic pursuit | Lower limbs | na | Y | N | N | na |
| N | N | N | N | N | N | N | N | na |
| N | N | N | N | N | N | N | N | N |
| N | Y | N | N | na | N | na | na | na |
| N | Y | N | N | Y | N | Y | Y | na |
| N | Y | N | N | Y | N | N | Y | na |
| Y | N | Supranuclear | N | na | N | N | Y | na |
| Y | N | Vertical | N | na | Y | na | na | na |

| VAS-spectrum related clinical impairment |  |  | Imaging |  |
| --- | --- | --- | --- | --- |
| Chronic cough | Neuropathic pain | NCS | Abnormal DaTSCAN | Brain MRI |
| *na* | **Y** | *na* | *na* | Cerebellar and brainstem atrophy |
| *na* | N | **Sensory** | *na* | Cerebellar and brainstem atrophy |
| *na* | **Y** | **Sensory** | *na* | Cerebellar and brainstem atrophy |
| N | N | *na* | *na* | Normal |
| N | N | *na* | *na* | Normal |
| *na* | *na* | *na* | *na* | Normal |
| **Y** | **Y** | **Axonal sensory** | Y | Normal at 46-50 |
| *na* | **Y** | **Axonal sensory-motor** | *na* | Normal |
| **Y** | N | **Sensory** | Y | T2-hyposignal of the posterior part of the putamen |
| *na* | *na* | *na* | Y | *na* |

| Comments |
| --- |
| Onset at 41-45 with urinary dysfunction, ataxia at 46-50, Gastrostomy and tracheostomy at 56-60<br>Optic atrophy |
| Classic form of PD, severe deterioration of dysarthria, swallowing disorders and major postural instability<br>DBS at 61-65 |
| DBS at 56-60, dyarthria after DBS |
| Erectile dysfunction and urinary imperiosis at 46-50<br>Urinary incontinence and erectile dysfunction at 61-65, Duodopa pump and then orthostatic hypotension at 66- |

### Appendix 1. PREDISTIM Study group

Lille

- Neurologists: Dr Caroline Moreau, Pr Luc Defebvre, Dr Nicolas Carriere, Dr Guillaume Grolez, Dr Guillaume Baille, Dr Kreisler
- Neuroradiologists: Pr Jean-Pierre Pruvo, Pr Xavier Leclerc, Dr Renaud Lopes, Dr Romain Viard, Dr Gregory Kuchcinski, Mr Julien Dumont
- Neuropsychologists: Pr Kathy Dujardin, Mme M Delliaux, Mme M Brion, Mme Virginie Herlin
- Neurosurgeons: Dr Gustavo Touzet, Pr Nicolas Reyns
- Neurophysiologists: Pr Arnaud Delval
- Clinical Assistant: Mme Valerie Santraine, Mme Marie Pleuvret, Mme Nolwen Dautrevaux, Mr Victor Laugeais, Mme Morgane Coeffet
- Clinical trials vigilance unit: Thavarak Ouk, Camille Potey, Celine Leclercq, Elise Gers Paris
- Neurologists: Jean-Christophe Corvol, Marie-Vidailhet, Elodie Hainque, Marie-Laure Welter, Lucette Lacomblez, David Grabli, Emmanuel Roze, Yulia Worbe, Cécile Delorme, Hana You, Jonas Ihle, Raquel Guimeraes-Costa, Florence Cormier-Dequaire, Aurélie Méneret, Andréas Hartmann, Louise-Laure Mariani
- Neuroradiologists: Stéphane Lehericy
- Neuropsychologists: Virginie Czernecki, Fanny Pineau, Frédérique Bozon, Camille Huiban, Eve Benchetrit, Marie Alexandrine Glachant
- Neurosurgeons: Carine Karachi, Soledad Navarro, Philippe Cornu
- Clinical Assistant: Arlette Welaratne, Carole Dongmo-Kenfack
- Nurses: Lise Mantisi, Nathalie Jarry, Sophie Aix, Carine Lefort Nantes
- Neurologists: Dr Tiphaine Rouaud, Pr Philippe Damier, Pr Pascal Derkinderen, Dr Anne-Gaelle Corbille
- Neuroradiologists: Dr Elisabeth Calvier-Auffray
- Neuropsychologists: Madame Laetitia Rocher, Madame Anne-Laure Deruet
- Neurosurgeons: Dr Raoul Sylvie, Dr Roualdes Vincent
- Clinical Assistant: Mme Le Dily Séverine Clermont-Ferrand
- Neurologists: Dr Ana Marques, Dr Berangere Debilly, Pr Franck Durif, Dr Philippe Derost, Dr Charlotte Beal
- Neuroradiologists: Carine Chassain
- Neuropsychologists: Laure Delaby, Tiphaine Vidal
- Neurosurgeons: Pr jean Jeacques Lemaire
- Clinical Assistant: Isabelle Rieu, Elodie Durand Marseille
- Neurologists: Pr Alexandre Eusebio, Pr Jean-Philippe Azulay, Dr Tatiana Witjas, Dr Frédérique Fluchère, Dr Stephan Grimaldi
- Neuroradiologists: Pr Nadine Girard
- Neuropsychologists: Eve Benchetrit, Marie Delfini
- Neurosurgeons: Dr Romain Carron, Pr Jean Regis, Dr Giorgio Spatola
- Clinical Assistant: Camille Magnaudet Poitiers
- Neurologists: Dr Ansquer Solène, Dr Benatru Isabelle, Dr Colin Olivier, Pr Houeto JL
- Neuroradiologists: Pr Guillevin Remy
- Neuropsychologists: Mme Fradet Anne, Mme Anziza Manssouri, Mme Blondeau Sophie
- Neuropsychiatrist: Dr Richard Philippe
- Neurosurgeons: Dr Cam Philippe, Dr Page Philippe, Pr Bataille Benoit
- Clinical Assistant: Mme Rabois Emilie, Mme Guillemain Annie Rennes
- Neurologists: Dr Drapier Sophie, Dr Frédérique Leh, Dr Alexandre Bonnet, Pr Marc Vérin
- Neuroradiologists: Dr Jean-Christophe Ferré
- Neuropsychologists: Mr Jean François Houvenaghel
- Neurosurgeons: Pr Claire Haegelen
- Clinical Assistant: Mme Francoise Kestens; Mme Solenn ory Bordeaux
- Neurologists: Pr Pierre Burbaud, Dr Nathalie Damon-Perriere, Pr Wassilios Meissner, Pr Francois Tison, Dr Stéphanie Bannier, Dr Elsa Krim, Pr Dominique Guehl
- Neuroradiologists: Sandrine Molinier-Blossier, Morgan Ollivier, Marion Lacoste
- Neuropsychologists: Nicolas Auzou, Marie Bonnet
- Neurosurgeons: Pr Emmanuel Cuny, Dr Julien Engelhardt
- Clinical Assistant: Olivier Branchard, Clotilde Huet, Julie Blanchard Toulouse
- Neurologists: Pr Rascol Olivier, Dr Christine Brefel Courbon, Dr Fabienne Ory Magne, Dr Marion Simonetta Moreau
- Psychiatric: Pr Christophe Arbus
- Neuroradioligst: Pr Fabrice Bonneville et Dr Jean Albert Lotterie
- Neuropsychologist: Marion Sarrail
- Neurosurgeon: Pr Patrick Chaynes, Pr François Caire
- Clinical Assistant: Estelle Harroch Rouen
- Neurologists: Pr David Maltete, Dr Romain Lefaucheur, Dr Damien Fetter
- Neuroradiologists: Dr Nicolas Magne
- Neuropsychologists: Mme Sandrine Bioux, Mme Maud Loubeyre, Mme Evangéline Bliaux, Mme Dorothée Pouliquen
- Neurosurgeon: Pr Stéphane Derrey
- Nurse: Mme Linda Vernon
- Biologist: Dr Frédéric Ziegler Strasbourg
- Neurologists: Mathieu Anheim, Ouhaid Lagha-Boukbiza, Christine Tranchant, Odile Gebus, Solveig Montaut
- Neuroradiologists: Stéphane Kremer
- Neuropsychologists: Nadine Longato, Clélie Phillips
- Neurosurgeons: Jimmy Voirin, Marie des Neiges Santin, Dominique Chaussemy
- Psychiatrist: Dr Amaury Mengin Nice
- Neurologists: Dr Caroline Girodana, Dr Claire Marsé
- Neuroradiologists: Lydiane Mondot
- Psychiatrics: Bruno Giordana, Robin Kardous
- Neuropsychologists: Bernadette Bailet, Héloise Joly
- Neurosurgeons: Denys Fontaine, Dr Aurélie Leplus
- IDE: Amélie Faustini
- Clinical Assistant: Vanessa Ferrier Amiens
- Neurologists: Pr Pierre Krystkowiak, Dr Mélissa Tir
- Neuroradiologists: Pr Jean-Marc Constans
- Neuropsychologists: Sandrine Wannepain
- Clinician Psychologist: Audrey Seling
- Neurosurgeon: Dr Michel Lefranc
- Clinical Assistant: Stéphanie Blin
- Parkinson coordinator IDE: Béatrice Schuler Lyon
- Neurologists: Pr Stephane Thobois, Dr Teodor Danaila, Dr Chloe Laurencin
- Neuroradiologists: Pr Yves Berthezene, Dr Roxana Ameli
- Neuropsychologists: Helene Klinger
- Neurosurgeons: Dr Gustavo Polo, Patrick Mertens
- Nurse: A Nunes
- Clinical Assistant: Elise Metereau Nancy
- Neurologists: Dr Lucie Hopes, Dr Solène Frismand
- Neuroradiologists: Dr Emmanuelle Schmitt
- Neuropsychologists: Mme Mylène Meyer, Mme Céline Dillier
- Neurosurgeon: Pr Sophie Colnat
- Clinical Assistant: Mme Anne Chatelain Hospital Fondation Rothschild
- Neurologists: Dr Jean-Philippe Brandel, Dr Cécile Hubsch, Dr Patte Karsenti, Dr Marie Lebouteux,Dr Marc Ziegler
- Neuroradiologists: Dr Christine Delmaire, Dr Julien Savatowky
- Neuropsychologists: Mme Juliette Vrillac, Mme Claire Nakache
- Neurosurgeon: Dr Vincent D’Hardemare
- Clinical Assistant: Mr Lhaouas Belamri Hospital Foch
- Neurologists: Dr Frédérique Bourdain, Dr Vadim Afanassiev, Dr Philippe Graveleau, Dr Cécilia Bonnet, Dr Valérie Mesnage, Dr Jarbas Correa Lino Junior
- Neurophysiologist: Dr Camille Decrocq
- Neuroradiologists: Dr Anne Boulin
- Neuropsychologists: Mme Elodie Dupuy, Mme Inès Barre
- Psychiatrics: Dr Bérénice Gardel
- Neurosurgeons: Pr Béchir Jarraya
- Clinical Assistant: Mme Delphine Lopez, Mr Christophe Fruit
- Coordinator: Mme Catherine Ziz

CATI (MRI acquisition management, preprocessing and data management)

David Gay, Robin Bonicel, Fouzia El Mountassir, Clara Fischer, Jean-François Mangin, Marie Chupin, Yann Cointepas

CRB of Lille (Center of Biological Resources)

Bertrand Accart, Patrick Gelé, Florine Fievet, Matthieu Chabel, Virginie Derenaucourt, Loïc Facon, Yanick

Tchantchou Njosse, Dominique Deplanque

Data management of Lille

Alain Duhamel, Lynda Djemmane, Florence Duflot, Emeline Cailliau

## Abbreviations

CANVAS: Cerebellar ataxia, neuropathy, and bilateral vestibular areflexia syndrome
CNIL: “Commission Nationale de l’Informatique et des Libertés”
DBS: deep brain stimulation
IQR: interquartile range
MSA: multiple system atrophy
MSA-c: multiple system atrophy of the cerebellar type
NCS: nerve conduction studies
PCR: polymerase chain reaction
PD: Parkinson’s disease
PREDI-STIM: Predictive Factors for therapeutic response on quality of life of Subthalamic Stimulation in Parkinson’s disease study
PSP: progressive supranuclear palsy
RFC1: replication factor C subunit 1
RP-PCR: repeat-primed polymerase chain reaction

## Authorship

### Authors

**Delforge V:** Major role in the acquisition of data, analysis or interpretation of the data, drafting or revising the manuscript for intellectual content

**Coarelli G:** Major role in the acquisition of data, drafting or revising the manuscript for intellectual content

**Mutez E:** Major role in the acquisition of data, analysis or interpretation of the data, drafting or revising the manuscript for intellectual content

**Rolland A-S:** Major role in the acquisition of data, drafting or revising the manuscript for intellectual content

**Wissocq A:** Major role in the acquisition of data, drafting or revising the manuscript for intellectual content

**Labudeck A:** Major role in the acquisition of data, drafting or revising the manuscript for intellectual content

**Marzys C:** Major role in the acquisition of data, drafting or revising the manuscript for intellectual content

**Boucetta N:** Major role in the acquisition of data, drafting or revising the manuscript for intellectual content

**Buée L:** Drafting or revising the manuscript for intellectual content

**Blum D:** Drafting or revising the manuscript for intellectual content

**Marques A-R:** Major role in the acquisition of data, drafting or revising the manuscript for intellectual content

**Demurger F:** Major role in the acquisition of data, drafting or revising the manuscript for intellectual content

**Azulay J-P:** Major role in the acquisition of data, drafting or revising the manuscript for intellectual content

**Lanore A:** Major role in the acquisition of data, drafting or revising the manuscript for intellectual content

**Tesson C:** Major role in the acquisition of data, drafting or revising the manuscript for intellectual content

**Lesage S:** Drafting or revising the manuscript for intellectual content

**Brice A:** Drafting or revising the manuscript for intellectual content

**Grabli D:** Major role in the acquisition of data, drafting or revising the manuscript for intellectual content

**Devos D:** Major role in the acquisition of data, drafting or revising the manuscript for intellectual content

**Durr A:** Major role in the acquisition of data, drafting or revising the manuscript for intellectual content

**Huin V:** Design or conceptualization of the study, major role in the acquisition of data, analysis or interpretation of the data, drafting or revising the manuscript for intellectual content

### Coinvestigator

PREDISTIM Study group

