## Supplementary figures and images for "Parkinson’s disease and multiple system atrophy are gateways to *RFC1*-related disorders"

### Supplementary Figure S1

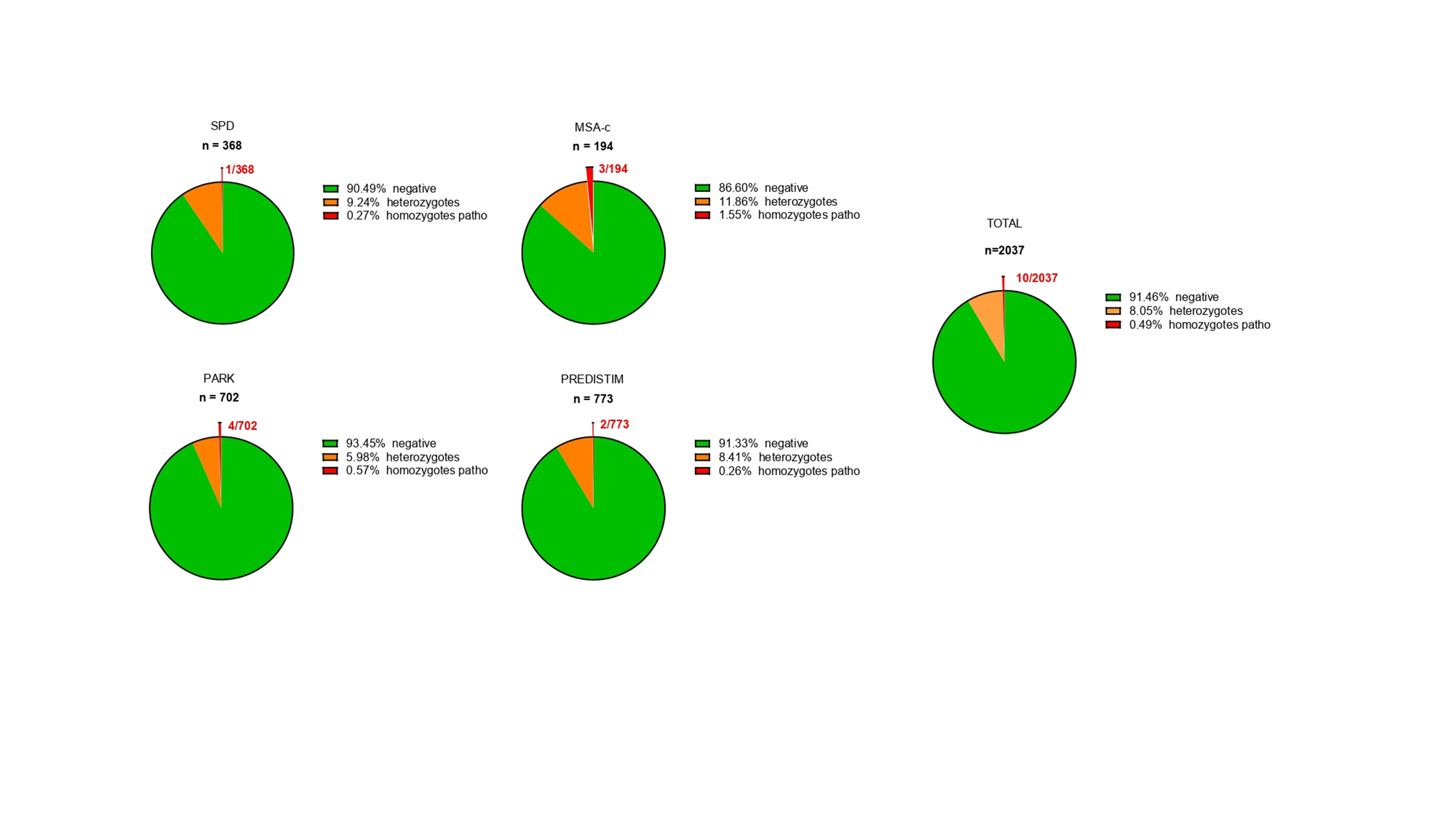
